# Epidemiological characteristics of patients with residual SARS-Cov-2 in Linyi, China

**DOI:** 10.1101/2020.06.16.20133199

**Authors:** Qiang Pan, Feng Gao, Rensheng Peng, Mingshan Li

**Affiliations:** Linyi Center for Disease Control and Prevention, Linyi, Shandong; Linyi Peoples Hospital, Linyi, Shandong; Center for Disease Control and Prevention of Hedong District, Linyi, Shandong; Fourth affiliated hospital of China Medical University, Shenyang, Liaoning

## Abstract

Patients with 2019 novel Coronavirus infection are probably show positive testing results again. In order to better treat these patients and provide basis for further control measures, we analyze the epidemiological outcomes and clinical features of patients with residual Severe Acute Respiratory Syndrome Coronavirus 2 (SARS-Cov-2) in Linyi city. From January 23 to March 31 in 2020, epidemiological and clinical information of confirmed patients are collected for analysis. Stool and pharyngeal swab samples are collected for RT-PCR testing. 64 confirmed patients are included and 17 patients present re-positive testing after discharge. For these 17 patients, 70.59% are family aggregated, the interval between first time of negative testing and first time of re-positive testing is 11.82±3.42 days. There is no difference between patients with continued negative testing results and re-positive testing. After discharge, the interval between first time of negative testing and first time of re-positive testing is associated with severity of disease (p=0.013). Besides, the duration from first time to last time of re-positive testing is associated with exposure or contact history (p=0.049) and severity of disease (p=0.001). The analysis reveals epidemiological characteristics of patients with residual SARS-Cov-2 and provide basis for further control measures.

## Introduction

From December in past year, 2019 novel Coronavirus (2019-nCov) started to spread around the world, causing more than 7.11 million cases of infection and 406 thousand cases of death. The virus transmits from people to people, and causes severe acute respiratory disease^1^. Linyi city locates in south of Shandong Province in the east of China, with a population of 11.25 million. By March 31 in 2020, 64 patients have been confirmed in Linyi, while 17 patients are positive again in RT-PCR testing for Severe Acute Respiratory Syndrome Coronavirus 2 (SARS-Cov-2) after discharge. Recently studies revealed that residual SARS-Cov-2 in lung tissues was the main reason for re-positive testing^2^. Thus these patients should be taken seriously. In order to better treat these patients with residual SARS-Cov-2 and provide the basis for further control measures, we analyzed the epidemiological outcomes and clinical features of 64 confirmed patients especially these 17 patients with residual SARS-Cov-2.

## Methods

### Participants and data collection

64 patients confirmed by laboratory testing for 2019-nCoV are included in the study. All the patients discharge with at least two continued negative testing for SARS-Cov-2, then they are centralized isolation for observation. The following testing during isolation is performed by the Linyi People’s Hospital or Linyi Centers for Disease Control and Prevention. The study ranges from January 23 to March 31 in 2020. Gender, age, exposure history to Hubei Province or contact with confirmed patients, onsets of symptoms, data of diagnosis, date of testing first negative, date of re-positive testing, severity of disease and other information were included for further analysis. All data were checked by two researchers (Q P and F G). The diagnosis, treatment and isolation for Coronavirus Disease 2019 (COVID-19) is according to the Chinese management guideline for COVID-19 (version 5.0)^3^. This study is approved by institutional review board of Linyi People’s Hospital. The need for informed consent is waived.

### Sample testing

Both stool and pharyngeal swab samples are collected for testing. The interval between two samples collection is not fixed. Real-time RT-PCR method^4^ was used for laboratory confirmation of SARS-CoV-2 infection.

### Statistical analysis

Chi-square test and ANOVA test were used to compare the difference where appropriate. A two-sided α of less than 0.05 was considered statistically significant. SPSS 22.0 was used for statistical analysis.

## Results

### Characteristics of confirmed patients

Among these 64 patients, 38 patients (59.37%) are male and 26 patients (40.63%) are female. The mean age is 39.76±17.89 years. 29 patients (45.31%) have exposure history of Hubei Province and 35 patients (54.62%) contact with confirmed patients or people return from Hubei Province. 15 patients (23.44%) have no symptom, 39 patients (60.94%) have symptoms of fever, cough or fatigue, 10 patients (15.62%) are severe in symptom and sign. 40 patients (62.5%) are family aggregated (Supplementary Table 1).

### Characteristics of patients with re-positive testing

For these patients with re-positive testing, 6 patients (35.29%) are female and 11 patients (64.71%) are male. The mean age is 37.37±21.57 years. 8 patients (47.06%) have exposure history of Hubei Province and 9 patients (52.94%) contact with confirmed patients or people return from Hubei Province. 4 patients are asymptomatic, 3 patients are mild, 8 patients are common and 2 patients are severe in illness. 1 case has a poor basic health state and low immunity (Supplementary Table 2). Among these patients, 2 patients are positive both in stool and pharyngeal swab, other patients are positive in stool samples. 9 patients are positive once again, 6 patients are positive twice again and 2 patients are positive thrice again. The interval between first time of negative testing and first time of re-positive testing is 11.82±3.42 days. After discharge, the duration of re-positive testing from the first time to the last time is 22.44±13.61 days (Figure 1). 12 patients (70.59%) are family aggregated, including a four-people family returned from Hubei Province with 3 members positive again and a big family with 6 confirmed patients. The second family extensive interact with each other during spring festival after one member contact with a confirmed patient, 5 people of the second family are positive again in following testing.

**Figure 1.**
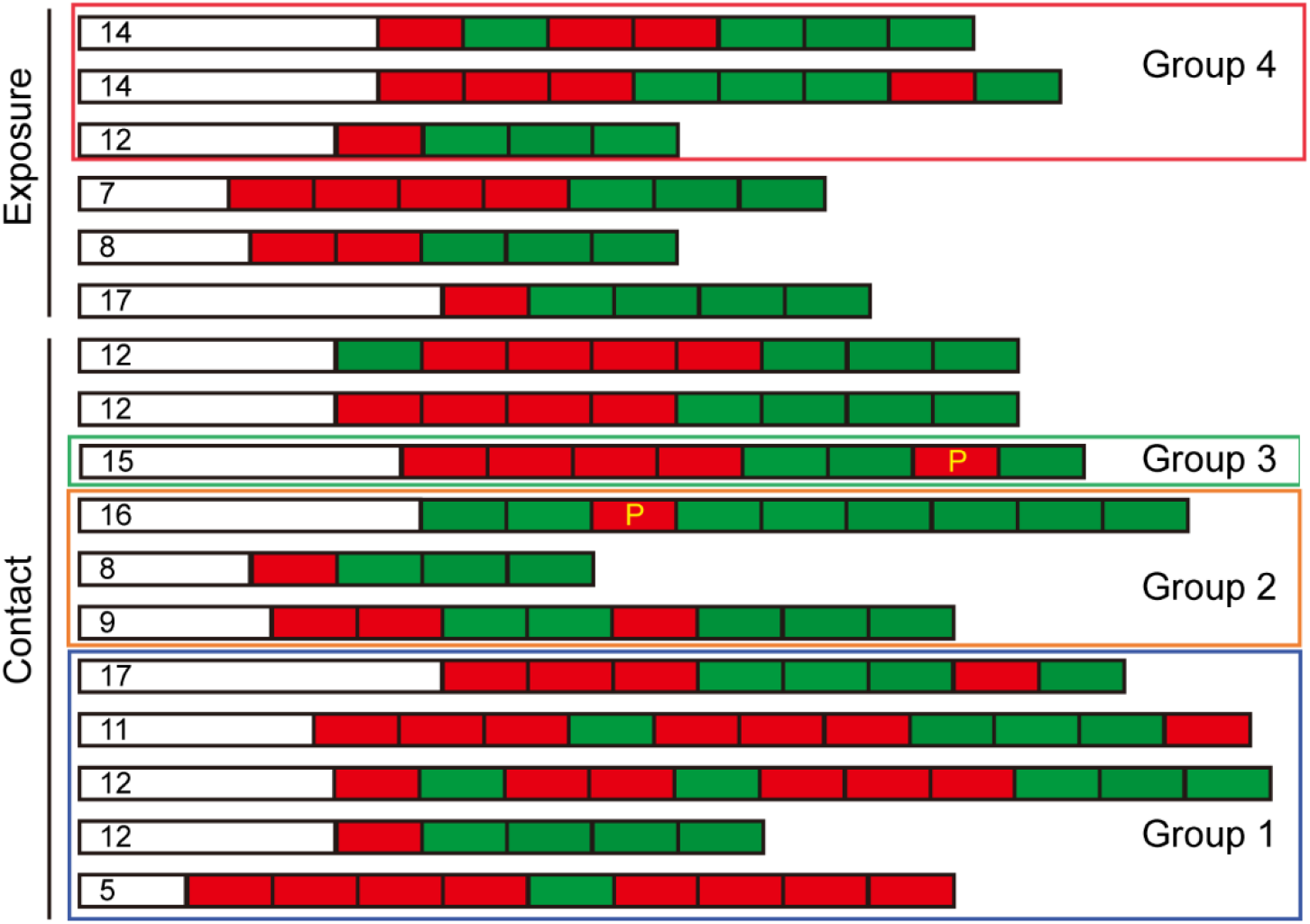
Epidemiological characteristics of 17 patients with re-positive testing. White bars refer to interval between first time of negative testing and first time of re-positive testing, the detail days are labelled; red bars refer to positive testing; green bars refer to negative testing. Family aggregation is labeled with detailed groups. P refers to samples are collected from pharyngeal swab, others are collected from stool.

### No difference between patients with negative testing and re-positive testing

To reveal whether patients with re-positive testing have particularity, we compare the features between patients with re-positive testing and continued negative. We find there is no difference in age (p=0.609), gender (p=0.602), severity (p=0.602), exposure to Hubei Province or contact with confirmed patients (p=0.333), aggregation (p=0.422). For patients with re-positive testing, the positive duration between onsets of symptoms and last positive testing in hospital is 16.29±5.58, while for patients with continued negative testing after discharge is 15.36±5.16 (p=0.541). There is no difference between patients with testing negative and re-positive testing (Supplementary Table 1).

### Relationship between clinical features and re-positive testing

Interval between first time of negative testing and first time of re-positive testing as well as the duration of re-positive testing from the first time to the last time are important monitoring indicators for confirmed patients. According to the analysis of patients with re-positive testing, we find age (p=0.346), gender (p=0.232), exposure or contact history (p=0.661) and aggregation (p=0.819) are not associated with the interval. Severity of disease (p=0.013) is significantly associated with the interval and the difference was mainly in comparison between slight symptom and serious illness (Asymptomatic+Mild vs Severe+Critic, p=0.005). Besides, we find age (p=0.95), gender (p=0.496), aggregation (p=0.186) are not associated with positive duration after discharge. Exposure or contact history (p=0.049) and severity of disease (p=0.019) are significantly associated with the positive duration (Table 1), which means that patients with serious illness have a longer time in duration of re-positive testing.

**Table 1.** Relationship between clinical features and positive outcomes

|  | Patients | Interval (days) | P value | Duration (days) | P value |
| --- | --- | --- | --- | --- | --- |
| Age |  |  | 0.346 |  | 0.95 |
| ≥60 | 1 | 8 |  | 25 |  |
| <60 and ≥18 | 11 | 11.18±3.71 |  | 21.64±14.91 |  |
| <18 | 4 | 13.25±1.50 |  | 24.00±15.45 |  |
| Gender |  |  | 0.232 |  | 0.496 |
| Male | 10 | 10.70±3.53 |  | 20.50±16.05 |  |
| Female | 6 | 12.83±2.85 |  | 25.67±10.42 |  |
| Severity |  |  | **0.013\*** |  | **0.019\*\*** |
| Asymptomatic<br>+Mild | 7 | 13.57±1.62 |  | 22.14±12.76 |  |
| Common | 7 | 10.59±3.34 |  | 14.00±7.35 |  |
| Severe+Critic | 2 | 6.50±2.12 |  | 40.00±13.45 |  |
| History |  |  | 0.661 |  | **0.049** |
| Exposure | 6 | 12.00±3.85 |  | 13.67±9.40 |  |
| Contact | 10 | 11.20±3.22 |  | 27.70±14.07 |  |
| Aggregation |  |  | 0.819 |  | 0.186 |
| Yes | 11 | 11.64±3.26 |  | 25.64±15.69 |  |
| No | 5 | 11.20±3.96 |  | 15.40±5.90 |  |

Interval refers to range between first time of negative testing and first time of re-positive testing.
Duration refers to range between first time and last time of re-positive testing after discharge.
\* P value for Asymptomatic+Mild vs Severe+Critic is 0.005;
\*\* P value for Asymptomatic+Mild vs Severe+Critic is 0.037, for Common vs Severe+Critic is 0.006.

## Discussion

This study focuses on 2019-nCoV infection in a city with a large population. We found there is no difference in the gender, age and exposure between patients with re-positive testing and testing negative after discharge. This analysis indicated that population with residual SARS-Cov-2 are not special group. However, we find more than 70% patients are family aggregated. Along with previous study^2^, we deduce that contact control may effectively reduce exposure to SARS-Cov-2 and reduce the chance of re-positive testing. In our study, we find that patients with serious illness appear positive testing earlier. The reason maybe that serious illness resulted from more virus infections, and more residual SARS-Cov-2 remained in the body of these patients, which makes the positive testing appear easier and more likely. This reason is similar with previous report^5^. The analysis also indicates that even discharge from hospital with negative testing at least two times, some patients still carry residual SARS-Cov-2 and show repeatedly positive testing outcome for 22.44±13.61 days. For these patients, three weeks or longer time of isolation and continuous detection for SARS-Cov-2 after discharge is valuable^6^.

There are some limitations in this analysis. Our analysis mainly study the epidemiological and clinical characteristics of patients in Linyi city, the study is regional and crowd limited. The duration of exposure or contact and the severity of imported patients from other cities are not detailed recorded. The testing collection is not performed day by day, which causes deviation to accurate calculation of interval and duration. In our study, we find patients with history of exposure to Hubei Province show a shorter day in positive duration after discharge. This means that exposure to Hubei is not necessarily infect more virus than contact with confirmed patients. Incomplete recording information maybe the reason for the difference in exposure or contact history. Besides, one case in the analysis is special. He is 62 years old, has a poor basic health state and low immunity. Considering that the poor immunity affect the response of immune system to virus infection, he is excluded for this analysis. Thus, only one case older than 60 years is included in the study, leading limitation to age comparison.

## Conclusion

Most patients with re-positive testing are family aggregated, more than three week isolation and observation are necessary for them. Patients with serious illness are earlier to appear re-positive testing, and have a longer time in positive testing for SARS-Cov-2 after discharge. The analysis reveals epidemiological and clinical characteristics of patients with residual SARS-Cov-2 and provide basis for further control measures.

## Data Availability

All data is available in the manuscript

## Contributions

Data collection: Q. P., F. G and R. P.; data analysis: Q. P. and M. L.; manuscript drafting: Q. P. and M. L.; manuscript revision: F. G and M.L. All authors reviewed the manuscript and approved the final version.

---

**Supplementary Table 1.** Difference between patients with re-positive and negative testing

|  | Patients | Outcome |  | P value |
| --- | --- | --- | --- | --- |
|  |  | Re-positive | Negative |  |
| All patients | 64 | 17 | 48 |  |
| Age |  |  |  | 0.609 |
| ≥60 | 9 | 2 | 8 |  |
| <60 | 54 | 15 | 39 |  |
| Gender |  |  |  | 0.602 |
| Male | 38 | 11 | 27 |  |
| Female | 26 | 6 | 20 |  |
| Severity |  |  |  | 0.602 |
| Asymptomatic | 15 | 4 | 11 |  |
| Mild | 7 | 3 | 4 |  |
| Common | 32 | 7 | 25 |  |
| Severe | 6 | 1 | 5 |  |
| Critic | 4 | 2 | 2 |  |
| History |  |  |  | 0.333 |
| Exposure | 29 | 6 | 23 |  |
| Contact | 35 | 11 | 24 |  |
| Aggregation |  |  |  | 0.422 |
| Yes | 40 | 12 | 28 |  |
| No | 24 | 5 | 19 |  |
| Positive duration<br>(days) |  | 16.29±5.58 | 15.36±5.16 | 0.541 |

Positive duration refer to range from onsets of symptom or to last time of positive testing, for asymptomatic patients, the range is from first positive laboratory testing to last time of positive testing.

**Supplementary Table 2.**
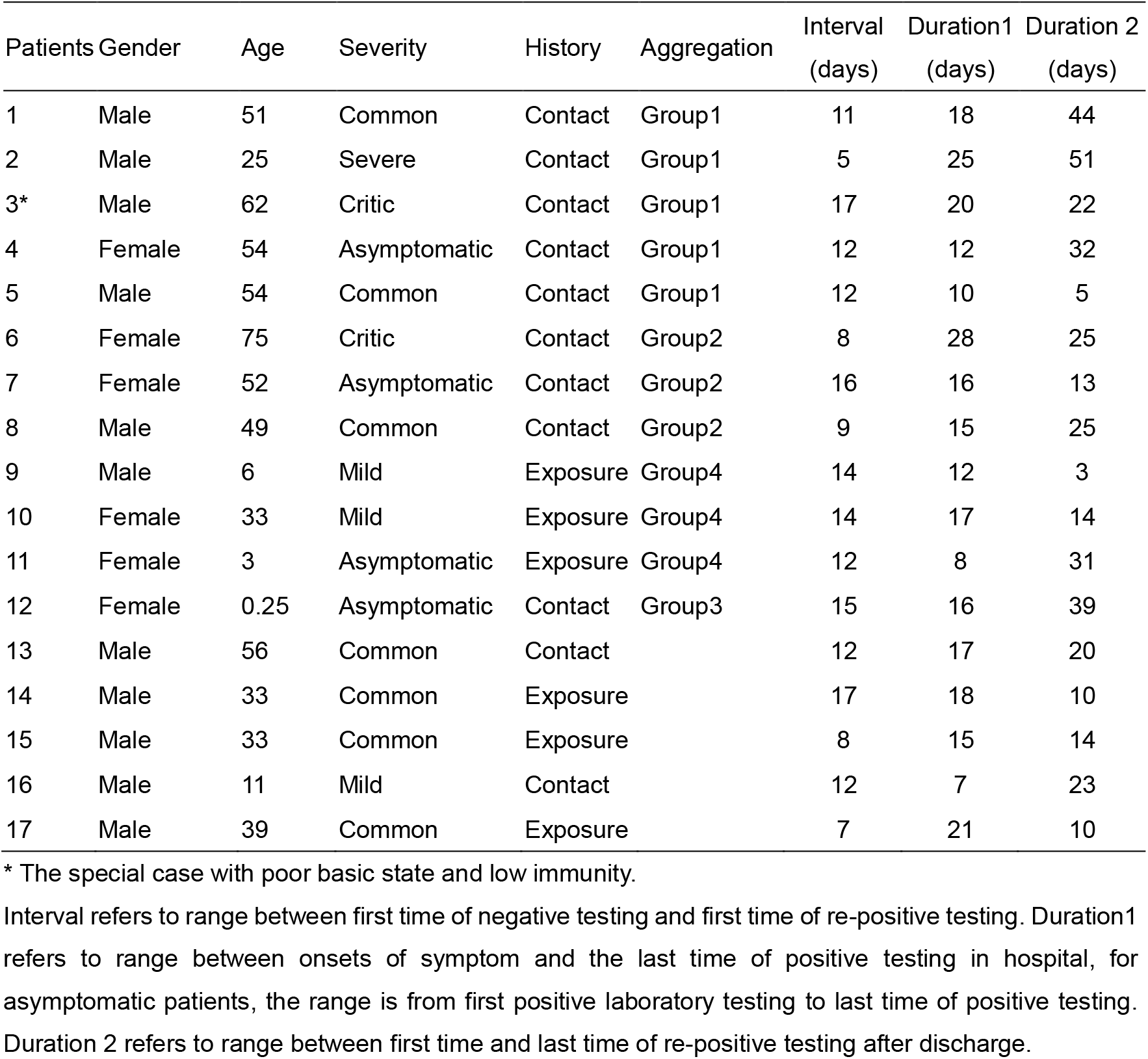
Characteristics of patients with re-positive testing.

| Patients | Gender | Age | Severity | History | Aggregation | Interval<br>(days) | Duration1<br>(days) | Duration 2<br>(days) |
| --- | --- | --- | --- | --- | --- | --- | --- | --- |
| 1 | Male | 51 | Common | Contact | Group1 | 11 | 18 | 44 |
| 2 | Male | 25 | Severe | Contact | Group1 | 5 | 25 | 51 |
| 3* | Male | 62 | Critic | Contact | Group1 | 17 | 20 | 22 |
| 4 | Female | 54 | Asymptomatic | Contact | Group1 | 12 | 12 | 32 |
| 5 | Male | 54 | Common | Contact | Group1 | 12 | 10 | 5 |
| 6 | Female | 75 | Critic | Contact | Group2 | 8 | 28 | 25 |
| 7 | Female | 52 | Asymptomatic | Contact | Group2 | 16 | 16 | 13 |
| 8 | Male | 49 | Common | Contact | Group2 | 9 | 15 | 25 |
| 9 | Male | 6 | Mild | Exposure | Group4 | 14 | 12 | 3 |
| 10 | Female | 33 | Mild | Exposure | Group4 | 14 | 17 | 14 |
| 11 | Female | 3 | Asymptomatic | Exposure | Group4 | 12 | 8 | 31 |
| 12 | Female | 0.25 | Asymptomatic | Contact | Group3 | 15 | 16 | 39 |
| 13 | Male | 56 | Common | Contact |  | 12 | 17 | 20 |
| 14 | Male | 33 | Common | Exposure |  | 17 | 18 | 10 |
| 15 | Male | 33 | Common | Exposure |  | 8 | 15 | 14 |
| 16 | Male | 11 | Mild | Contact |  | 12 | 7 | 23 |
| 17 | Male | 39 | Common | Exposure |  | 7 | 21 | 10 |

\* The special case with poor basic state and low immunity.
Interval refers to range between first time of negative testing and first time of re-positive testing. Duration1 refers to range between onsets of symptom and the last time of positive testing in hospital, for asymptomatic patients, the range is from first positive laboratory testing to last time of positive testing. Duration 2 refers to range between first time and last time of re-positive testing after discharge.

